# Population serum proteomics uncovers prognostic protein classifier and molecular mechanisms for metabolic syndrome

**DOI:** 10.1101/2022.10.21.22281353

**Authors:** Xue Cai, Zhangzhi Xue, Fang-Fang Zeng, Jun Tang, Liang Yue, Bo Wang, Weigang Ge, Yuting Xie, Zelei Miao, Wanglong Gou, Yuanqing Fu, Sainan Li, Jinlong Gao, Menglei Shuai, Ke Zhang, Fengzhe Xu, Yunyi Tian, Nan Xiang, Yan Zhou, Peng-Fei Shan, Yi Zhu, Yu-ming Chen, Ju-Sheng Zheng, Tiannan Guo

**Author notes:** Correspondence (Y.M.C); (J.S.Z.); (T.G.). These authors contributed equally (Xue Cai, Zhangzhi Xue, Fang-Fang Zeng, Jun Tang, Liang Yue, Bo Wang).

## Abstract

Metabolic syndrome (MetS) is a complex metabolic disorder with a global prevalence of 20-25%. Early identification and intervention would help minimize the global burden on healthcare systems. Here, we measured over 400 proteins from ∼20,000 proteomes using data-independent acquisition mass spectrometry for 7890 serum samples from a longitudinal cohort of 3840 participants with two follow-up time points over ten years. We then built a machine learning model for predicting the risk of developing MetS within ten years. Our model, composed of 11 proteins and the age of the individuals, achieved an area under the curve of 0.784 in the discovery cohort (n=855) and 0.774 in the validation cohort (n=242). Using linear mixed models, we found that apolipoproteins, immune-related proteins, and coagulation-related proteins best correlated with MetS development. This population-scale proteomics study broadens our understanding of MetS, and may guide the development of prevention and targeted therapies for MetS.

## INTRODUCTION

Metabolic syndrome (MetS), also called insulin resistance syndrome [1], is a complex metabolic disorder characterized by abdominal obesity, atherogenic dyslipidemia, raised blood pressure, insulin resistance, central obesity, prothrombotic and proinflammatory states [2, 3]. About 20-25% of adults worldwide suffer from MetS [4-6]. In Chinese adults, the prevalence of MetS reached 24.2% (24.6% in men and 23.8% in women) in 2018 [7]. According to national health and nutrition examination survey (NHANES), about one-third of US adults were diagnosed with MetS between 1988 and 2010 [8]. MetS has been reported to predispose to several serious diseases [9-11], including diabetes, cardiovascular disease, coronary heart disease, and several common cancers. An early prediction and diagnosis of MetS would allow faster interventions, reducing the burden on the healthcare systems. There are currently diagnostic criteria of MetS, with the main components of waist circumference, glucose, blood pressure, triglycerides (TG), high-density lipoprotein cholesterol (HDL-C) [3]. But the pathogenesis of MetS is unclear and there is still a lack of targeted therapy for MetS. Thus, longitudinal large cohort population studies are necessary, which can effectively predict the development and explore the pathogenesis of MetS.

Plasma and serum are the predominant samples used for diagnostic analyses in clinics with low invasiveness and ease of collection and preservation. Plasma and serum proteomic studies based on large cohort population have used aptamer-based (SOMAscan) platform [12] and proximity extension assay (Olink proteomics) [13] as tools for metabolic disease [14, 15]. For example, using the SOMAscan platform, Ganz et al quantified the plasma proteins of two longitudinal cohorts of 938 and 971 samples with one follow-up point, and achieved 0.70 of C statistics in validation cohort for cardiovascular risk prediction based on 9-protein panel [16]. Proteomics of 3301 human plasma samples from two different subcohorts were also measured by the SOMAscan platform to explore the non-linear alterations in the human plasma proteome with age [17]. Furthermore, dozens of blood proteins were measured by the proximity extension assay (Olink proteomics) to investigate the association with BMI and waist circumference in a cross-sectional study of 3,308 participants [18] and predict myocardial infarction risk with C-index of 0.68 in 5,131 diabetes patients [19].

The rapid development of mass spectrometry (MS)-based proteomics provided solid technical support needed by large cohort proteomics [20]. Specifically, this technology provides high-throughput and reproducible analyses with minimal sample amounts. Data-independent acquisition (DIA) MS measures all fliable peptide precursor ions without prior knowledge, allowing the unbiased discovery of new biomarkers [21, 22]. For instance, using DIA MS methods, Niu *et al*. analyzed the plasma samples of 596 individuals and 79 liver biopsies in three weeks and predicted future liver-related events and all-cause fatalities [23]. Bruderer *et al*. also analyze 1508 plasma samples with robust capillary-flow DIA MS to investigate potential biomarkers for wight loss [24].

Here, we reliably measured over 400 proteins from ∼20,000 proteomes using DIA-MS for 7890 samples from a longitudinal cohort of 3840 participants with two follow-up time points over ten years. Based on our data, we built a model to predict the risk of developing MetS within ten years. We also explored new potential biomarkers and networks associated with MetS, providing reference for the pathogenesis and targeted therapy of MetS.

## RESULTS

### Serum proteome profiling of a longitudinal cohort

Here, we included 3840 participants from the community-based prospective cohort study, namely Guangzhou Nutrition and Health Study (GNHS, ClinicalTrials.gov identifier: NCT03179657) [25-28]. Serum samples were collected at three of four time points: 3479 samples were collected at baseline (between 2008 and 2010), 2638 at the second follow-up (between 2014 and 2017), and 1773 at the third follow-up (between 2018 and 2019) (**Table 1**). No serum samples were collected at first follow-up. On average, 5.6 years passed between the baseline and the second follow-up, and 3.4 years between the second and the third follow-up. The statistics of the participants at the three time points are summarized in **Table 1**; their clinical information is provided in **Table S1**. In particular, the median age for the baseline, second follow-up, and third follow-up groups were 57.4 (38.2-80.4), 63 (44.3-83.3), and 66.1 (46.8-86.2), with 69%, 68%, and 69% female participants, respectively. We next divided the serum samples into a discovery (n=4794) and a validation cohort (n=3094) randomly (**Figure 1**), and the sample preparation, MS data collection, data analyses were then independently performed.

**Table 1.** Statistics of the 3,840 participants from the Guangzhou Nutrition and Health Study (GNHS).

|  | Baseline<br>(N=3480) | Second follow-up<br>(N=2639) | Third follow-up<br>(N=1773) |
| --- | --- | --- | --- |
| Age (years) | 57.4 (38.2-80.4) | 63 (44.3-83.3) | 66.1 (46.8-86.2) |
| Sex (% of women) | 69 | 68 | 69 |
| BMI (kg/m<sup>2</sup>) | 23.2 (14.8-51.3) | 23.5 (14.5-57.3) | 23.6(15.3-54.1) |
| Waist (men, cm) | 86.4 (61-147) | 87.8 (64.4-155.8) | 88 (64.5-162) |
| Waist (women, cm) | 81.2 (58-117.9) | 86.5 (60.3-128,6) | 83.6 (60.2-113.8) |
| Fasting serum glucose (mmol/L) | 4.7 (0.2-24.6) | 5 (3.2-24.6) | 5.3 (3.9-21.1) |
| Blood triglycerides (mmol/L) | 1.3 (0.01-25.4) | 1.3 (0.2-15.6) | 1.4 (0.45-15) |
| Serum HDL (mmol/L) | 1.4 (0.02-3.3) | 1.5 (0.6-4.6) | 1.4 (0.6-4.3) |
| Serum LDL (mmol/L) | 3.6 (0.1-10) | 3.6 (0.6-11.5) | 3.5 (0.9-8) |
| SBP | 122 (73-228) | 122 (63.5-241) | 120 (76-205.7) |
| DBP | 79 (46-135) | 73.3 (43.7-120) | 72.5 (47.7-117.3) |
| SBP/DBP (mmHg) | 1.6 (1-2.7) | 1.6 (0.9-2.8) | 1.6 (1-2.5) |
| MetS (%) | 0.17 | 0.21 | 0.18 |

**Figure 1.**
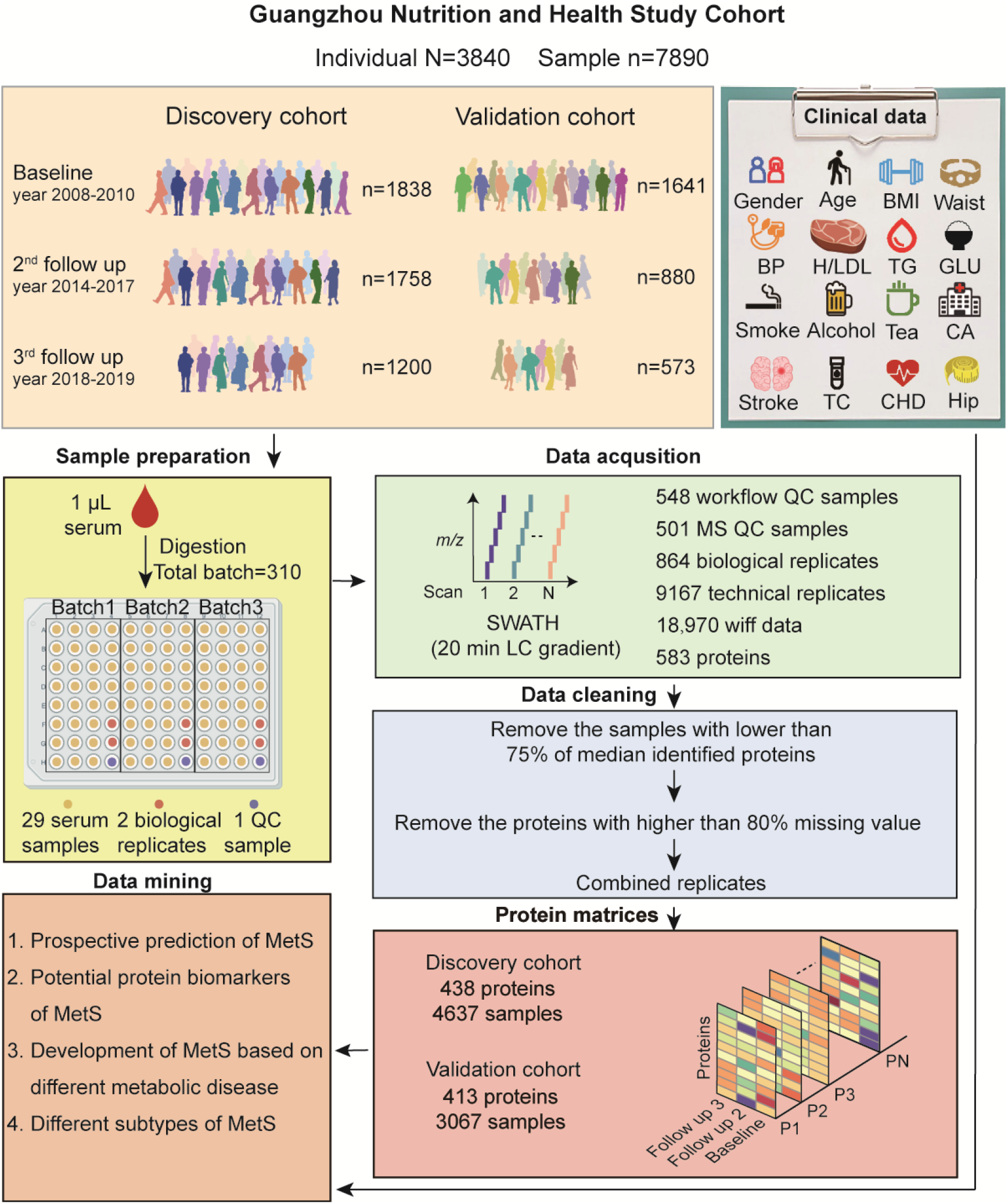
Study design. GNHS study population: 7890 samples (4796 in the discovery cohort, 3094 in the validation cohort) were collected from 3840 individuals. Altogether we generated 18,970 DIA-MS data. After data cleaning, the protein matrices (containing 4637 samples and 438 proteins for the discovery cohort; 3067 samples and 413 proteins for the validation cohort) were used for data mining and machine learning modeling.

We randomly divided the 4796 serum samples of our discovery cohort into 178 batches, each containing 29 unique serum samples, two biological replicates, and one quality control (QC) sample to ensure the stability of the entire workflow (**Figure 1, Table S2A**). An MS-QC sample was injected before each batch to ensure the instrument was in good condition. Next, each sample was acquired 2-3 times using a 20-min DIA-MS method [21], summing up to 11,646 MS data, including 301 QC samples, 526 biological replicates, 288 MS-QC samples, 5735 technical replicates (**Figure 1**). Finally, we used the software tool DIA-NN [29] to generate a matrix of 583 proteins and 11,646 MS data with 45% missing values.

To obtain high-quality proteomics data for our subsequent analyses, we cleaned and sorted the protein matrix returned by DIA-NN. First, we excluded 445 MS data with protein identifications below 75% of the median protein identification (<245), which may have been affected by an imperfect sample preparation or the instrument status. We also excluded proteins with missing values exceeding 80% of the protein matrix (n=145) (**Figure 1**). The median Pearson correlation coefficient (r) of the MS-QC samples achieved 0.94, proving the high stability of the MS instruments (**Figure 2A)**. Next, we checked the MS-QC samples of each batch and excluded 167 MS data with no MS-QC samples and no biological or technical replicates, as it would have been hard to assess the quality of their MS data. Finally, we corrected the batch effects using ProteomeExpert [30] and observed a significant improvement in our data (**Figure 2B)**. The Pearson correlation coefficients (r) of 512 biological replicates and 5317 technical replicates were calculated, and the median r of the two sets of replicates were 0.97 and 0.96 (**Figure 2C)**, respectively. These results prove the high consistency and reproducibility of our data. For our subsequent data analyses, we combined the quantitative results of those replicates with r >0.8.

**Figure 2.**
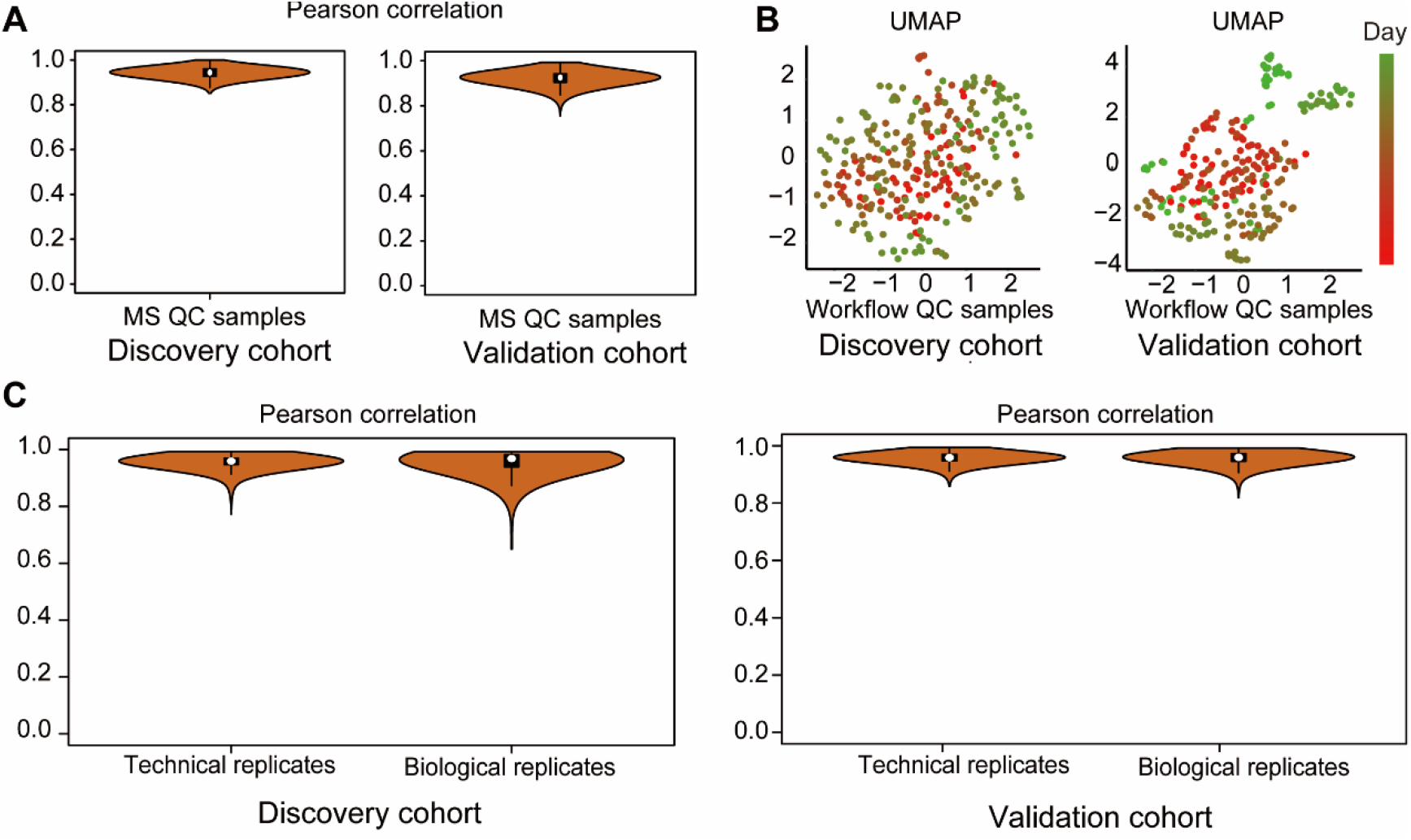
Quality control analyses and dataset pre-processing. **A)** Pearson correlation of the MS QC samples from the discovery and validation cohorts. **B)** Batch effect evaluation using PCA and the QC samples from the discovery and validation cohorts. **C)** Pearson correlation of the technical and biological replicates from the discovery and validation cohorts.

The experimental design of our validation cohort was the same as that of our discovery cohort. We divided 3094 serum samples into 132 batches (**Table S2A**). A total of 7324 MS data (containing 247 QC samples, 338 biological replicates, 213 MS-QC samples, and 3432 technical replicates) were measured using 20-min DIA-MS. Next, we excluded 194 MS data with protein identifications below 75% of the median protein identification (<238), and sub the protein matrix containing 413 proteins as we obtained from the discovery cohort. The median r values of the MS-QC samples, the biological replicates, and the technical replicates were 0.93 (**Figure 2A**), 0.96, and 0.95 (**Figure 2C**), respectively. We also corrected the batch effect using ProteomeExpert and observed a significant improvement (**Figure 2B**).

After these steps, from the discovery cohort we obtained a protein matrix containing 4637 samples and 438 proteins, with 19.0% missing values (**Figure 1, Table S2B**); from the validation cohort we obtained a protein matrix containing 3067 samples and 413 proteins, with 18.4% missing values (**Figure 1, Table S3B**).

### A protein-based classifier for predicting MetS insurgence

For the discovery cohort, we collected a set of samples at the baseline time point, including 267 non-MetS samples from individuals who were diagnosed with MetS at the second or third follow-up and 588 non-MetS samples from individuals who were not diagnosed with MetS at any of our time points (**Figure 3A**). Using these data, we built a machine learning model to evaluate the serum proteins’ ability to predict the risk of developing MetS within ten years.

**Figure 3.**
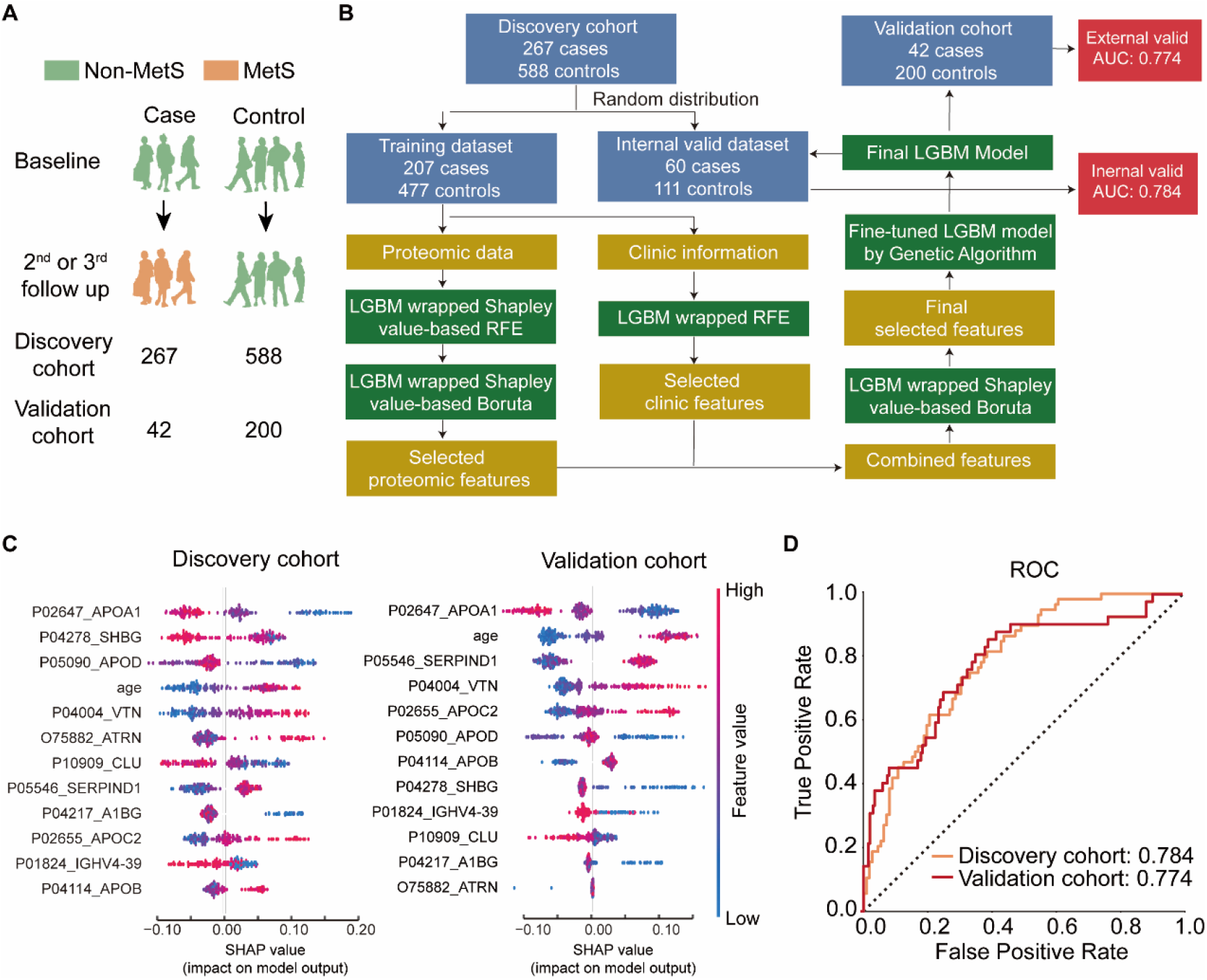
Machine learning model for predicting the risk of developing MetS. **A)** Case and control samples for the machine learning model. **B)** Workflow of the machine learning model built with quantitative proteomics data and clinical features. **C)** Shapley importance of variables in the model. **D)** Receiver operating characteristic (ROC) plot of the machine learning model.

We used LightGBM to generate a machine learning model using the discovery dataset of 855 samples containing 288 serum proteins, as well as age and sex information (**Figure 3B, Table S4A**). First, we randomly split the dataset into a training dataset (n=684) and an internal validation dataset (n=171). Then, a 2-step feature selection was used for choosing protein features from the 288 serum proteome (**Figure 3B**). Specifically, the final protein features were apolipoprotein A-I (APOA1), sex hormone-binding globulin (SHBG), apolipoprotein D (APOD), vitronectin (VTN), attractin (ATRN), clusterin (CLU), heparin cofactor 2 (SERPIND1, HCII), alpha-1B-glycoprotein (A1BG), apolipoprotein C-II (APOC2), immunoglobulin heavy variable 4-39 (IGHV4-39), and apolipoprotein B-100 (APOB) (**Figure 3C**). We optimized our model using a ten-fold cross-validation. Next, we tested the model using the randomly selected internal validation dataset and achieved an AUC of 0.78, indicating that our model can efficiently predict the risk of developing MetS within ten years (**Figure 3D**). Finally, we tested the model using 242 samples from an independent validation cohort and achieved an AUC of 0.77 (**Figure 3D, Table S4B**). These results show that the model generalized well to independently collected samples and the protein used in the model may present as promising biomarker candidates.

### Circulating proteins regulated in MetS samples

To find which serum proteins are potentially affected by MetS, we used two linear mixed models to examine: (1) the relationship between protein level and MetS at the baseline; (2) the dynamic relative changes along the three time points. For the baseline analysis, we used 1736 baseline samples (309 MetS, 1427 non-MetS); for the dynamic analysis, we used 4561 samples from all three timepoints (986 MetS, 3575 non-MetS). With the baseline analysis, we found that 175 proteins were significantly dysregulated in the MetS compared with the non-MetS group (**Table S5A**), while 194 proteins were associated with the dynamic development of MetS during ten years (**Table S5B**). A total of 143 proteins were dysregulated according to both analyses (**Figure 4A**).

**Figure 4.**
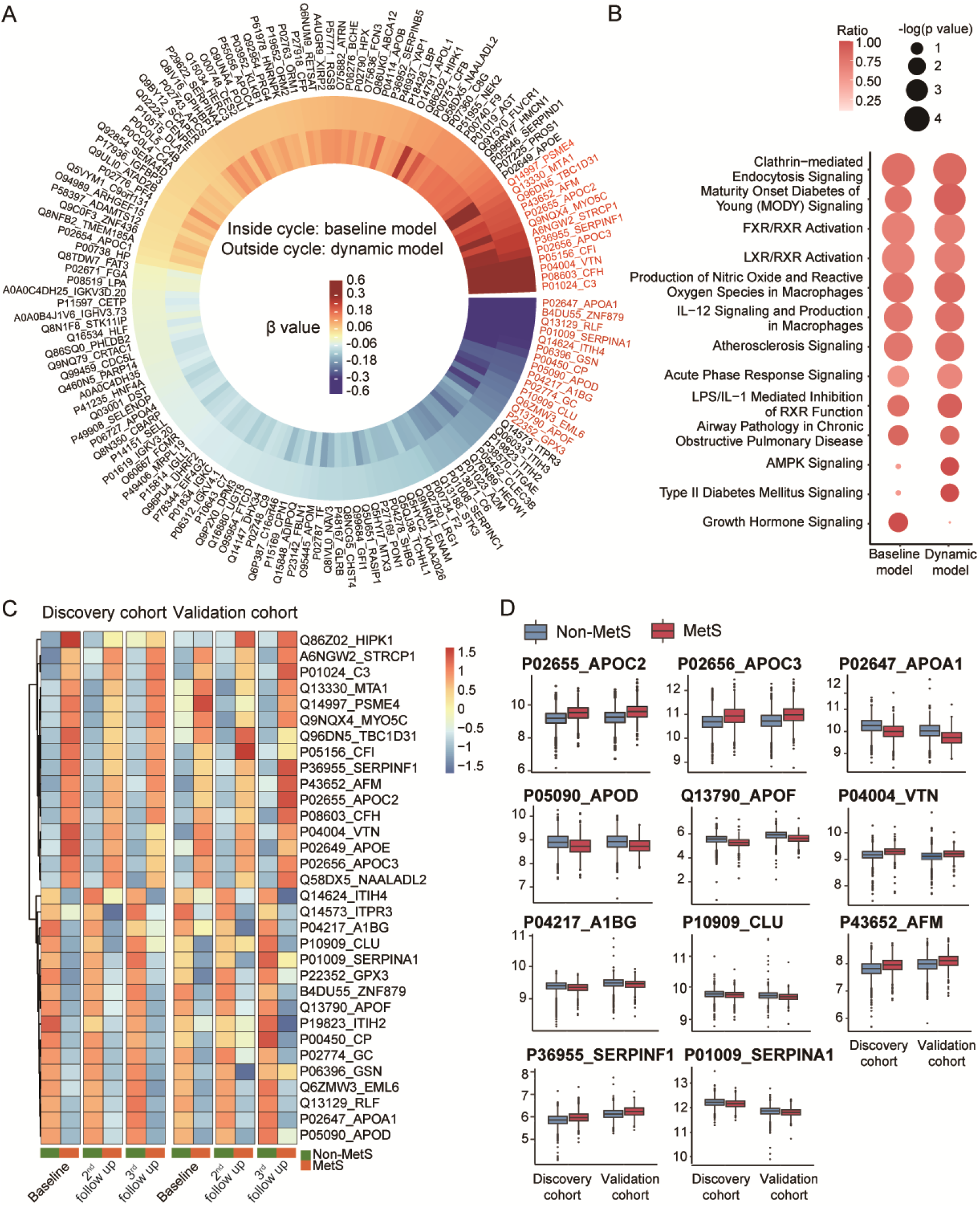
Performance of new potential protein biomarkers of MetS and their related pathways. **A)** Dysregulated proteins highly associated with MetS were selected using linear mixed models. **B)** Enriched pathways analyzed by IPA of 175 and 194 significantly dysregulated proteins. **C)** Top (|β|>0.2) significantly dysregulated proteins associated with MetS using the linear mixed model. **D)** Expression of typical dysregulated proteins in discovery cohort and validation cohort.

To find which pathways may be affected by MetS, we analyzed the 175 and 194 dysregulated proteins using Ingenuity Pathway Analysis (IPA). We found that 11 pathways were significantly dysregulated according to our baseline analysis (**Figure 4B, Table S5C**), and 12 were significantly dysregulated according to our dynamic analysis (the threshold for both analyses was -log(p-value)>1.3) (**Figure 4B, Table S5D**). In particular, ten pathways were significantly dysregulated according to both analyses, including FXR/RXR activation, LXR/RXR activation, atherosclerosis signaling, maturity-onset diabetes of young (MODY) signaling, and acute-phase response signaling (**Figure 4B**). Seven pathways with the most significant p-values were mainly enriched in apolipoproteins (**Table S5C, 5D**); among these seven, FXR/RXR activation and LXR/RXR activation are involved in metabolism and associated with RXR activation [31]. RXR activation is known to correlate highly with MetS, as drugs targeting RXR and its heterodimeric partners are already in clinical use against MetS [32]. Additionally, MetS has been reported to increase the risk of atherosclerosis [33] and is a clinical characteristic of MODY [34]. Acute-phase response signaling has also been associated with MetS [35]. The growth hormone signaling pathway was significantly dysregulated only in the baseline analysis (**Figure 4B**), indicating this pathway was dysregulated in MetS patients but possibly not linked with its development. In particular, the increase in growth hormone can cause insulin resistance in men [36]. Type II diabetes mellitus and the AMPK signaling pathways were significantly dysregulated in the dynamic analysis only (**Figure 4B**), indicating these pathways may be primarily involved in the development of MetS. Diabetes is a component of MetS [37], and evidence suggests that the dysregulation of the AMPK signaling pathway may lead to MetS [38].

The expression of 32 significantly dysregulated proteins (|β|>0.2, see Methods), among the 175 proteins and 194 proteins, showed the same performance in the discovery and the validation cohorts (**Figure 4C)**. These proteins were dysregulated in MetS with respect to the non-MetS patients and were linked to the development of MetS. The elevated expression of APOC2 and VTN (**Figure 4D**), and the lower expression of AOPA1, APOD, A1BG, CLU, and APOC2 (**Figure 4D**) in MetS patients were consistent with the results of above analysis (**Figure 3D**). In addition, two apolipoproteins, apolipoprotein C-III (APOC3) and apolipoprotein F (APOF), and several other reported proteins, pigment epithelium-derived factor (PEDF), alpha-1-antitrypsin (A1AT) and afamin (AFM) were also dysregulated in MetS patients (**Figure 4D**). These results show that apolipoproteins and their related pathways are closely related to the occurrence and development of MetS.

### Molecular characterization of MetS subtypes

MetS can be diagnosed when at least three of five diagnostic indicators are met: waist circumference, fasting blood glucose, blood pressure (SBP/DBP), fasting triglyceride (TG) and fasting high-density lipoprotein (HDL)-C (see Methods) [37]. Subtypes of MetS are based on different indicators. Here, we compared the different types of MetS and non-MetS and explored the molecular mechanism of MetS based on different indicators.

We selected 444 MetS samples that met the indicators of abnormal TG (**Table S6A, B**) and 444 non-MetS control samples with normal TG and matching age, sex, and other MetS indicators same as MetS. Similarly, we selected 169, 235, 384, and 354 paired samples to analyze abnormal HDL, glucose, waist, and SBP/DBP, respectively (**Table S6A, B**). No dysregulated proteins were discovered in the waist- and SBP/DBP-based MetS samples and only one up-regulated protein was deregulated in the glucose-based MetS (**Figure S1**). On the other hand, six up-regulated and three down-regulated proteins were observed in the TG-based MetS samples (**Figure 5A**), while one up-regulated and six down-regulated proteins were observed in the HDL-based MetS samples (**Figure 5B**). The expressions of these dysregulated proteins were consistent in the discovery and validation cohorts (**Figure 5C, 5D**).

**Figure 5.**
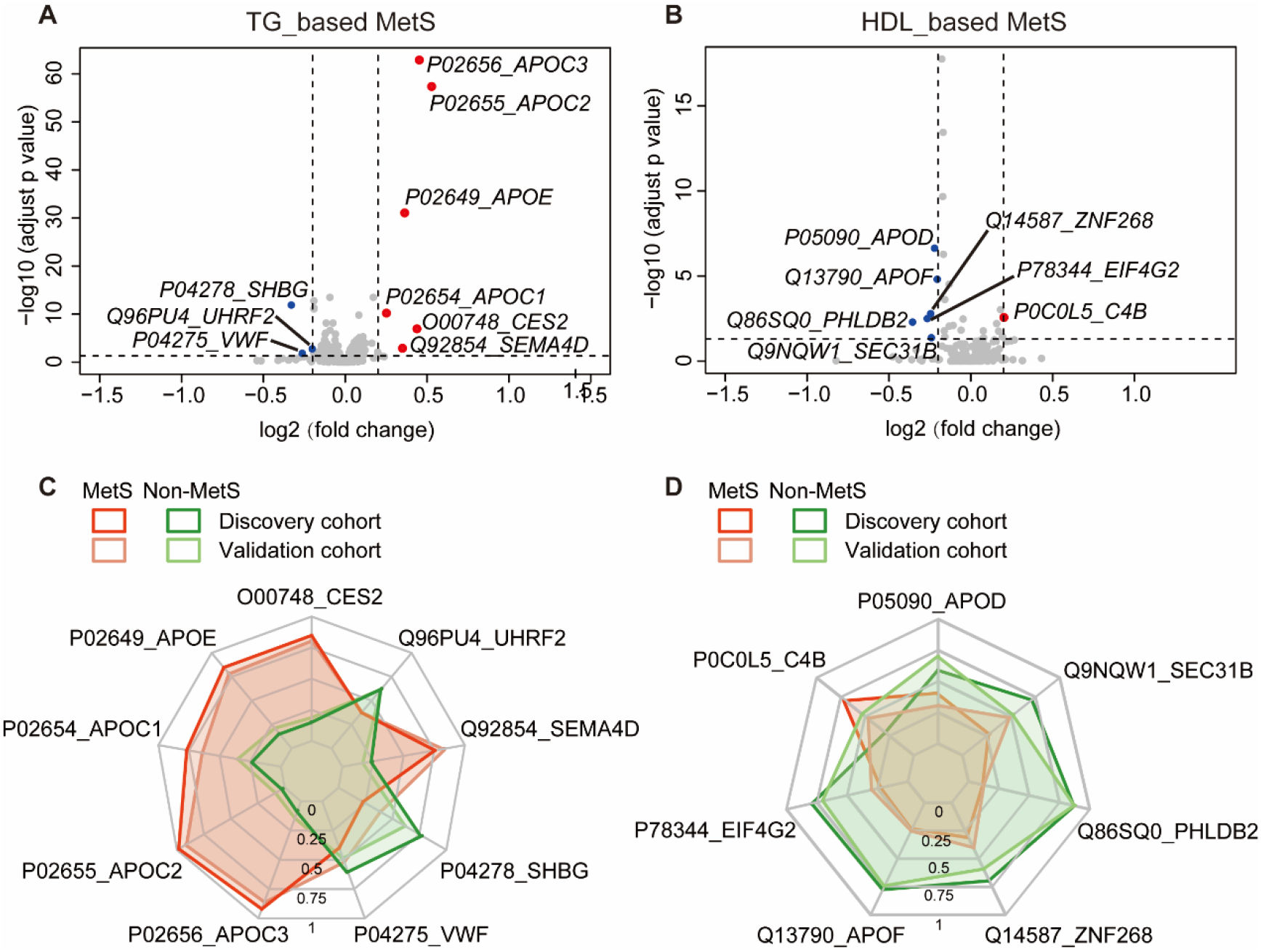
Dysregulated proteins in different types of MetS. **A)** Volcano plot of the dysregulated proteins in the TG-based MetS. **B**) Volcano plot of the dysregulated proteins in the HDL-based MetS. **C**) Expression of the dysregulated proteins in the TG-based MetS and the non-MetS. **D**) Expression of the dysregulated proteins in the TG-based MetS and the non-MetS.

Next, we analyzed the dysregulated proteins in TG- and HDL-based MetS samples using the IPA. The network enriched in the dysregulated proteins from the TG-based MetS samples (**Figure S2**) showed that the increased abundance of APOE, APOC1, APOC2, and APOC3 led to the activation of HDL and LDL, whereas predict the inhibition of VLDL-cholesterol and insulin. In contrast, in the HDL-based MetS, the decrease in APOD and APOF led to the inhibition of HDL, and predicted the inhibition of LDL (**Figure S3**).

### Proteins associated with MetS initiation

To investigate the development mechanisms of MetS, we next focused on participants that were not diagnosed with MetS at the baseline but developed MetS within ten years. By looking at the five diagnostic indicators of MetS mentioned above, some of these people met 1-2 indicators, others none. Therefore, we explored the proteins that significantly changed in the serum of participants that developed MetS based on the five abnormal indicators.

We selected 146 individuals from the discovery cohort that only met the waist indicator for MetS at baseline (**Table S7A, E**). Of these individuals, only 19 developed MetS. As the remaining 127 participants had an abnormal waist indicator also at the two follow-up visits, they acted as controls. To minimize the influence of sample size, we employed oversampling in our analysis (see Methods). In the MetS patients, 12 proteins were significantly up-regulated, while 24 were significantly down-regulated at the baseline compared with the control group (**Figure 6A, Table S7B**). These proteins included angiotensinogen (AGT), apolipoprotein D (APOD), and thrombospondin-1 (THBS1).

**Figure 6.**
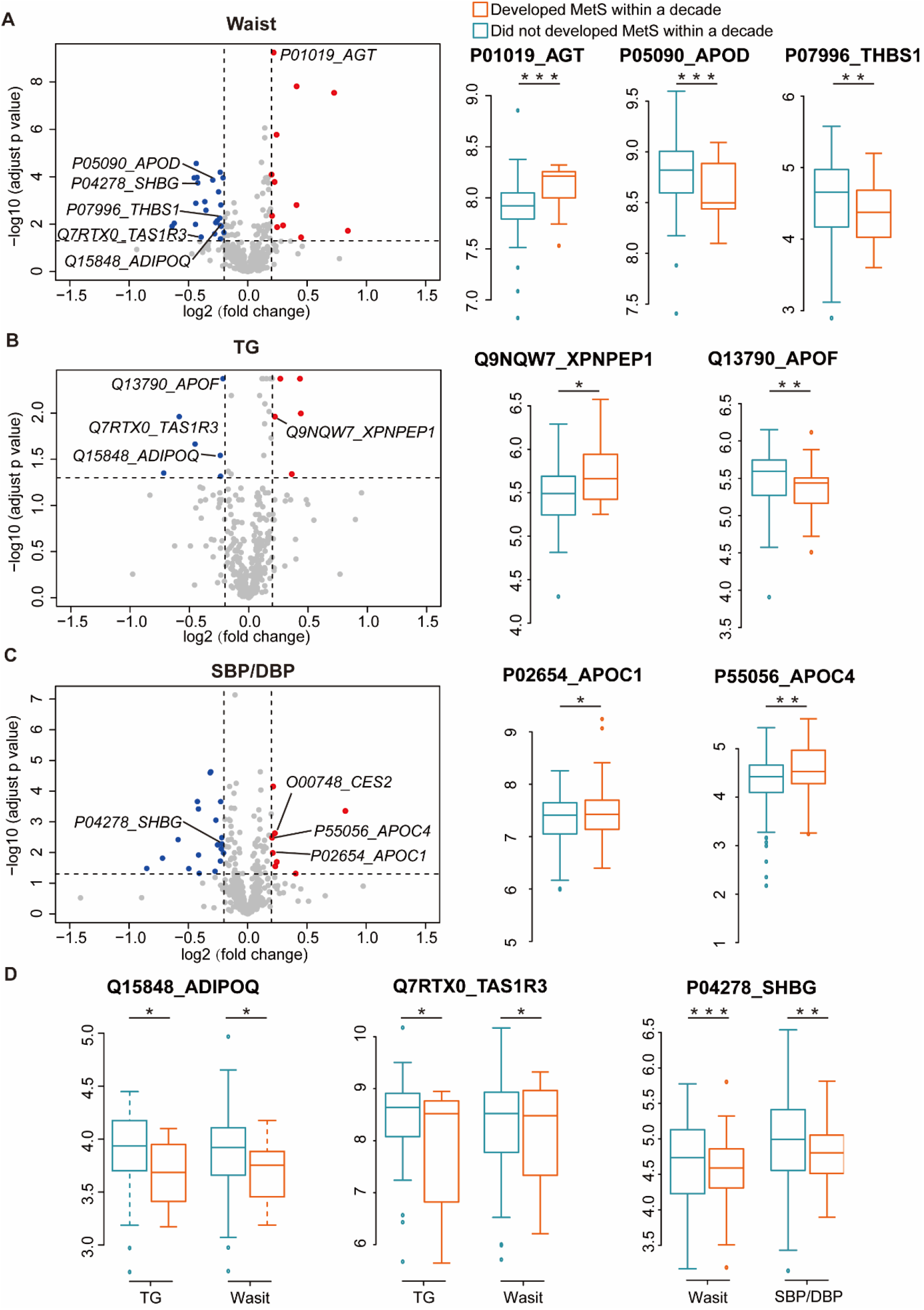
Dysregulated proteins in the development of MetS. **A**) Volcano plot and boxplots of the dysregulated proteins in the development of waist-based MetS. **B**) Volcano plot and boxplots of the dysregulated proteins in the development of TG-based MetS. **C**) Volcano plot and boxplots of the dysregulated proteins in the development of SBP/DBP-based MetS. **D**) Boxplots of the overlapping core dysregulated proteins in the development of waist-based MetS, TG-based MetS, and SBP/DBP-based MetS. Adjust p value: *, <0.05; **, <0.01; ***, <0.001.

We next focused on TG. A total of 24 cases and 112 controls were selected similarly to the previous analysis on the waist indicator. Only five proteins were significantly up-regulated, and six were significantly down-regulated (**Figure 6B, Table S7A, C**), including Xaa-Pro aminopeptidase 1 (XPNPEP1) and apolipoprotein F (APOF). Using the SBP/DBP indicator, 45 cases and 246 controls were selected; eight proteins were significantly up-regulated and 19 were significantly down-regulated (**Figure 6C, Table S7A, D**), including apolipoprotein C-I (APOC1) and apolipoprotein C-IV (APOC4). Finally, as less than ten cases satisfied the selection criteria based on the blood glucose and HDL indicators, we did not analyze them.

Among the dysregulated proteins we identified, adiponectin (ADIPOQ) and taste receptor type 1 member 3 (TAS1R3) were dysregulated both in the TG-based and the waist-based MetS analyses (**Figure 6D**). This finding suggests that these two proteins may play a key role in the development of MetS. Additionally, SHBG was dysregulated both in the TG-based and the SBP/DBP-based MetS analyses, and was also selected in our machine learning model for predicting MetS risk.

## DISCUSSION

We generated a population proteomics resource based on 7,890 serum samples collected from a prospective cohort of 3,840 participants over ten years, with a total of 18,970 DIA-MS data. We then generated a machine learning model to predict the risk for an individual of developing MetS over the next ten years. We also used this dataset to explore the molecular bases of MetS onset and development.

To our knowledge, our study provides the largest DIA-based serum proteome from a prospective population cohort with two follow-up time points to date. Niu *et al*. recently built a dataset of 311 plasma proteins from 596 individuals using a 21-min DIA-MS method and Orbitrap Exploris 480 mass spectrometer [23]. Also, Orwoll *et al*. identified 224 proteins in 1,196 serum samples using liquid chromatography-ion mobility-mass spectrometry (LC-IMS-MS) with a 58-min LC gradient [39]. In our study, using a 20-min DIA-MS method, 438 and 413 proteins were identified, with 19% and 18% missing values, in the independent protein matrices of the discovery and the validation cohorts, respectively.

A combination of blood proteomics and physiological status in a longitudinal population study can be used for disease risk prediction, as it has been shown for various metabolic diseases with high incidence [40, 41]. We used this method to study MetS. In particular, we built a prospective machine learning model to predict the risk of developing MetS within ten years.

Elhadad *et al*. generated a LASSO model for the prospective analysis of incident MetS (mean follow-up time = 6.5) in a German population based on the data of 8 proteins [15]. Their training cohort of 623 participants achieved an AUC of 0.74, but no validation cohort was used. Our study used 11 serum proteins plus age as model features. The AUC of our model was 0.784 in the discovery cohort (855 individuals) and 0.774 in the validation cohort (242 individuals). Also, our resource has an additional follow-up time point of 10 years, providing more accurate data for the long-term prediction of MetS.

Our proteomics analysis provides essential insights into MetS, highlighting the apolipoproteins that play a crucial role in its pathophysiology. MetS is also known as insulin resistance syndrome [1], and apolipoproteins changes are associated with insulin resistance [9]. In particular, APOC3, a component of the triglyceride-rich very low-density lipoproteins (VLDL) and HDL in plasma [42], had been reported as a MetS risk factor in the Canadian [43] and Turkish populations [44]. Our results showed that apolipoproteins, APOC2, APOC3, APOA1, APOD, and APOF are not only associated with MetS onset but also with the development of MetS. Also, the expression of different apolipoproteins showed important variations in our data: APOC2 and APOC3 had high expression levels, while APOA1, APOD, and APOF had low expression levels. The higher expressions of APOC1, APOC2, APOC3, and APOE were already reported in MetS [45]. Therefore, exploring the biology of different apolipoproteins can provide more precise knowledge of the pathophysiology of MetS. In our data, APOC1, APOC2, APOC3, and APOE were up-regulated in the TG-based MetS, while APOD and APOF were down-regulated in HDL-based MetS. These observations suggest that apolipoproteins may have different roles and influences in MetS of different subtypes. Indeed, APOCs play an essential role in the etiology of human hyperlipidemias [46], especially APOC2 [47] and APOC3 [48]. Our data showed that these proteins may be related to MetS primarily by affecting TG. Circulating APOD and APOF are mainly present in HDL [49, 50] and may thus be related to MetS by affecting HDL.

In addition to apolipoproteins, we found that several other proteins are significantly associated with MetS development. Some of these proteins were already linked to MetS by other studies. SHBG can regulate the plasma metabolic clearance rate of steroid hormones and, in agreement with our findings, has been reported to predict the development of MetS in a population-based cohort study of middle-aged Finnish men [51]. Vitronectin is a multifunctional adhesive glycoprotein that exerts regulatory functions in coagulation, fibrinolysis, or the plasminogen activation system [52]. In a prospective study, baseline plasma vitronectin was found as a marker of incident MetS at nine years [53], consistently with our findings. Also, the serum levels of PEDF encoded by SERPINF1 (a glycoprotein that belongs to the superfamily of serine protease inhibitors) have been reported associated with MetS in a cross-sectional study of the Japanese population [54]. Furthermore, in a 10-year prospective study of Chinese men, PEDF was also associated with the development of the MetS [55]. Alpha-1-antitrypsin (A1AT) encoded by SERPINA1, is an acute-phase inflammation marker that is associated with the development of the MetS [56]. AFM, a human plasma vitamin E-binding glycoprotein, was found to be strongly associated with the prevalence in a cross-sectional manner and development of MetS in a prospectively epidemiological study [57]. We also identified proteins that have not been reported to be directly associated with MetS. A1BG, belongs to the immunoglobulin family, was reported to be elevated in the urine of diabetes patients [58, 59]. Interestingly, the lower expression was observed in the serum of patients with MetS or those who develop MetS within 10 years, indicating that the protein may be related to the humoral regulatory system of MetS. Another immune-related protein, attractin, shows a higher level in circulating blood monocytes of human subjects because of obesity [60]. IGHV4-39, an immunoglobulin produced by B lymphocytes, has been reported to promote insulin resistance [61]. These results suggest that the immune system and coagulation related systems may also be important regulatory systems in the development of MetS.

The core mechanism of metabolic diseases, such as hypertension, hyperglycemia, hyperlipidemia, and obesity, which often appear together and develop into MetS, is still unclear. Here we explored the dysregulated proteins linked to the development of MetS. We found that specific dysregulated proteins were connected with various metabolic abnormal states and MetS development, providing a new basis for understanding this mechanism. Among these proteins, we found AGT, THBS1, XPNPEP1, adiponectin, and TAS1R3. AGT is an essential component of the renin-angiotensin system (RAS), and increased insulin stimulates AGT production [62]. We found a higher expression of AGT in the population that later developed MetS, suggesting that AGT positively correlates with the development of MetS consistently with other studies [63]. THBS1, an adipose-derived matricellular protein, has been reported as a biomarker of MetS in Japanese subjects [64]. Adiponectin is an anti-inflammatory cytokine and a biomarker for the development of MetS [9, 65], and it was downregulated in our data. A lower adiponectin level was observed in Chinese adolescents [66], peri- and post-menopausal women [67], young Polish subjects [68] with MetS, and in a prospective study of the Korean General Population with MetS [69]. Finally, TAS1R3 is a putative taste receptor reported to negatively correlate with glucose levels, triglycerides, and MetS [70]. XPNPEP1 contributes to the degradation of bradykinin, and the bradykinin system has beneficial effects on hypertension [71, 72] and type 2 diabetes [73]. This agrees with our data showing higher levels of XPNPEP1 in the populations that later developed MetS. These findings are again consistent with our results and corroborate the robustness of our findings, which include other proteins that have not yet been linked to MetS development.

The findings of this study have to be seen in the light of some limitations. First, we used a short gradient, high-throughput proteomics workflow, which is more suitable for analyzing large cohorts. Consequently, the serum proteins we identified are relatively high in abundance. Also, our discovery and validation cohort are from the same city. Our results, therefore, require a follow-up study using independent populations from different areas.

The GNHS community-based prospective cohort study was initiated in 2008. Comprehensive phenotypic data for the individuals involved in this study have been recorded over time, including but not limited to questionnaire, physical examination, dual-energy x-ray absorptiometry, ultrasonography, and blood/urine/faces tests [25-28]. The proteomic data acquired in this study could be potentially applied in understanding other health status and diseases such as diabetes.

In conclusion, we generated a resource of serum proteomics using DIA-MS based on a prospective population cohort with a 10-year follow-up. Using this data, we built a model for predicting the risk of developing MetS within ten years. We also found new potential protein biomarkers of MetS that, together with their pathways, providing new perspectives on the pathophysiology of MetS.

## MATERIALS AND METHODS

### Patients and samples

This study was based on the previously reported community-based prospective cohort Guangzhou Nutrition and Health Study (GNHS, ClinicalTrials.gov identifier: NCT03179657) [25-28]. Briefly, 3840 of 4048 participants aged 40-83 were enrolled in this study. The participants were recruited between 2008 and 2013 and followed up until 2019 (**Figure 1, Table 1, Table S1A**). In particular, participants were evaluated and blood samples were taken from them at up to three time points: baseline (between 2008 and 2013), second follow-up (between 2014 and 2017), and third follow-up (between 2018 and 2019). Venous whole blood samples were collected from all participants early in the morning before having food using serum separation tubes, and then centrifuged at 3500 rpm for 10 min for serum collection. The serum samples were frozen at −80 °C before the analysis.

The diagnostic criteria for MetS in this study refer to the 2016 Chinese guidelines for managing dyslipidemia in adults [37]. Specifically, they included three or more of the following: central obesity/abdominal obesity: a waist circumference of at least 90 cm for men and at least 85 cm for women; hyperglycemia: fasting blood glucose of at least 6.10 mmol/L (110 mg/dL) or 2-h blood glucose after glycemic load of at least 7.80 mmol/L (140 mg/dL) or confirmed diabetes for patients that received treatment; hypertension: blood pressure of at least 130/85 mmHg or patients with hypertension that received treatment; fasting TG of at least 1.7 mmol/L (150 mg/dL); fasting HDL-C lower than 1.0 mmol/L (40 mg/dL).

### Protein digestion

Peptides were extracted from the serum samples as previously described [74, 75]. Briefly, 1 μL of serum sample was lysed using 20 μL of lysis buffer (8 M urea (Sigma, Catalog # U1230) in 100 mM ammonium bicarbonate, ABB) at 32° C for 30 min, then reduced and alkylated using 10 mM tris (2-carboxyethyl) phosphine (Sigma Catalog # T4708) and 40 mM iodoacetamide (Sigma, Catalog # SLCD4031), respectively. Before the enzymatic digestion, 70 μL of 100 mM ABB were added to the samples to dilute the urea. The protein extracts were then digested with a two-step overnight tryptic digestion (Hualishi Tech. Ltd, Beijing, China), using an enzyme-to-substrate ratio of 1:60 (final ratio 1:30) at 32° C for 4 h and then another 12 h. The digestion was then stopped by adjusting the pH to 2-3 using 1% trifluoroacetic acid (Thermo Fisher Scientific, Catalog # T/3258/PB05). Before the MS analysis, peptides were cleaned with HRP SOLAu columns (Thermo Fisher Scientific™, San Jose, USA).

### Mass spectrometric analysis

The peptide samples were injected into an Eksigent NanoLC 400 System (Eksigent, Dublin, CA, USA) coupled with a TripleTOF 5600 system (SCIEX, CA, USA) for the SWATH-MS analysis, as previously described [75]. Briefly, 0.5 μg of peptides were separated with a 20-min LC gradient of 5-30% buffer B. Buffer A contained 2% acetonitrile (ACN) and 0.1% formic acid (FA) in HPLC water, while buffer B contained 98% ACN and 0.1% FA in HPLC water. For SWATH-MS, a 55 variable Q1 isolation window scheme was set as in previous studies [75].

### Proteome data analysis

The MS files were analyzed using DIA-NN (1.8) [76] against a plasma spectral library [75] containing 5102 peptides and 819 unique proteins from the Swiss-Prot database of *Homo sapiens*. In the DIA-NN settings, the software automatically sets the retention time extraction window, while the *m/z* extraction window for MS1 and MS2 was set to 20 ppm and 50 ppm, respectively. Protein and peptide false discovery rates were set not to exceed 1%. Protein inference was set to the protein names (from the FASTA file), and the cross-run normalization was set as ‘RT-dependent’.

### Machine learning

In the machine learning analysis, we used LightGBM Library (Ver. 3.3.2) in Python (Ver. 3.9) to generate a machine learning model using the discovery dataset of 855 samples containing 288 serum proteins, as well as age and sex information. First, we randomly split the dataset into a training dataset (n=684) and an internal validation dataset (n=171). Then, a 2-step feature selection was used for choosing protein features from the discovery dataset and other features from the patient’s clinical information (*i*.*e*. age, gender) to be included in the final classifier. The features were firstly selected based on proteome data and clinical data independently. In the second selection step, preselected features of the two categories were combined and any unrelated proteome was further eliminated, especially when the proteome was correlated with age and sex. The purpose of splitting the feature selection into two steps was to preserve proteomes that may have a weak contribution to classification, as including clinical information in the first place may result in the elimination of these proteomes due to their stronger contributions. During the first step, we used a Shapley value-based recursive feature elimination (RFE) algorithm, followed by a Shapely value-based Boruta algorithm for serum proteome feature selection, and the feature importance-based RFE for age and gender selection. In the second step, we combined preselected proteomes and clinical information from the first step and performed another Shapley value-based Boruta algorithm to obtain the final feature set. Through feature selection, 11 proteins and age were selected as the final features. We then optimized our model using the training dataset with ten-fold cross-validation and tested the model with the internal validation dataset and the independent validation cohort.

### Statistical analysis

The linear mixed model for the baseline analysis used the data collected at the baseline visit, while the linear mixed model for the dynamic analysis used the data collected from all three time points. For the two linear mixed models, MetS and non-MetS were set as outcome variables; the protein expression, age, and sex were set as fixed effects; the patient ID was set as a random effect in the linear mixed model for dynamic analysis. The value β represents the regression coefficient, which indicates the influence of independent variable effect on dependent outcome variable in the regression equation. The larger the regression coefficient is, the greater the influence of effect on outcome variable is. The positive regression coefficient means that outcome variable increases with the increase of effect, while the negative regression coefficient means that outcome variable decreases with the increase of effect.

## Data Availability

All the raw data will be publicly released after publication. The phenotype data can be requested by email from the corresponding author (Y.C).

## ACKNOWLEDGMENTS

This work is supported by grants from the National Key R&D Program of China (No. 2021YFA1301602, 2021YFA1301603, and 2021YFA1301601), the National Natural Science Foundation of China (No. 82073546, 81773416, and 82103826) and Zhejiang Provincial Natural Science Foundation of China (LQ21H260002). We thank Westlake University Supercomputer Center for assistance in data generation and storage. We thank all study participants of the Guangzhou Nutrition and Health Study and all other team members involved in the cohort study.

## AUTHOR CONTRIBUTIONS

T.G., J.Z, Y.C., and Y.Z. designed and supervised the project. Y.C. and F.Z. established the cohort and collected the phenotype data and the samples. X.C., J.T., L.Y., Y.X., Z.M., W.G., Y.F., S.L., M.S., K.Z., F.X., Y.T., N.X., and Y.Z. generated the data. X.C., Z.X., B.W., W.G., and J.G. analyzed the data. X.C., L.Y., P.S., Y.Z., and T.G. drafted the manuscript with inputs from all co-authors. X.C., Z.X, F.Z., J.T., L.Y., and B.W. contribute equally to this work. We thank Z.R., Y.L., H.C., Z.L., B.W., and H.M for assistance in data generation and analysis.

## SUPPLEMENTAL INFORMATION

Table S1. Clinical information of the study participants from our GNHS cohort.

Table S2. Proteomics data of the discovery cohort.

Table S3. Proteomics data of the validation cohort.

Table S4. Proteomics and clinical data for our machine learning modeling.

Table S5. Proteins associated with MetS and its development.

Table S6. Proteomics data for the analysis of different MetS subtypes.

Table S7. Proteomics data for the analysis of MetS development.

**Figure S1.**
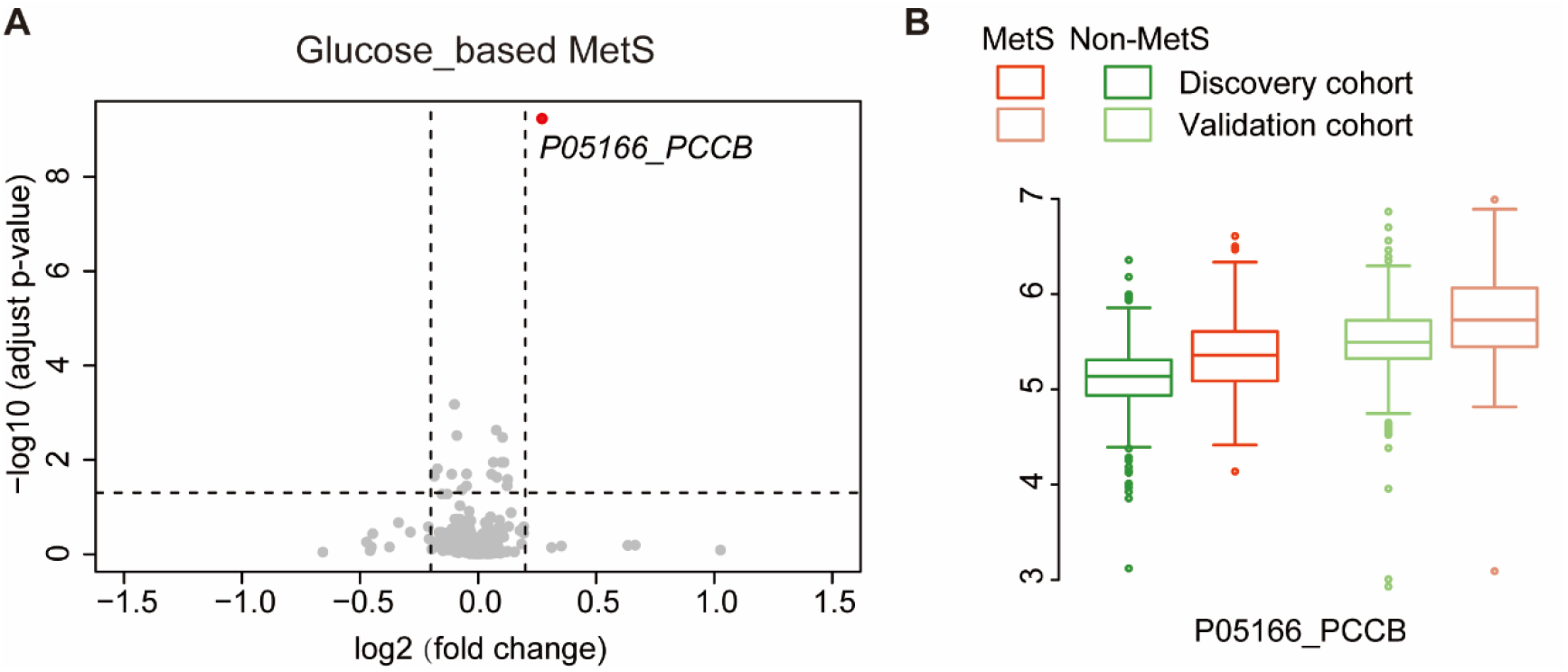
Dysregulated proteins from the glucose-based MetS patients. **A**) Volcano plot of the dysregulated proteins from the glucose-based MetS patients. **B**) Expression of the dysregulated proteins from the glucose-based MetS and the non-MetS patients.

**Figure S2.**
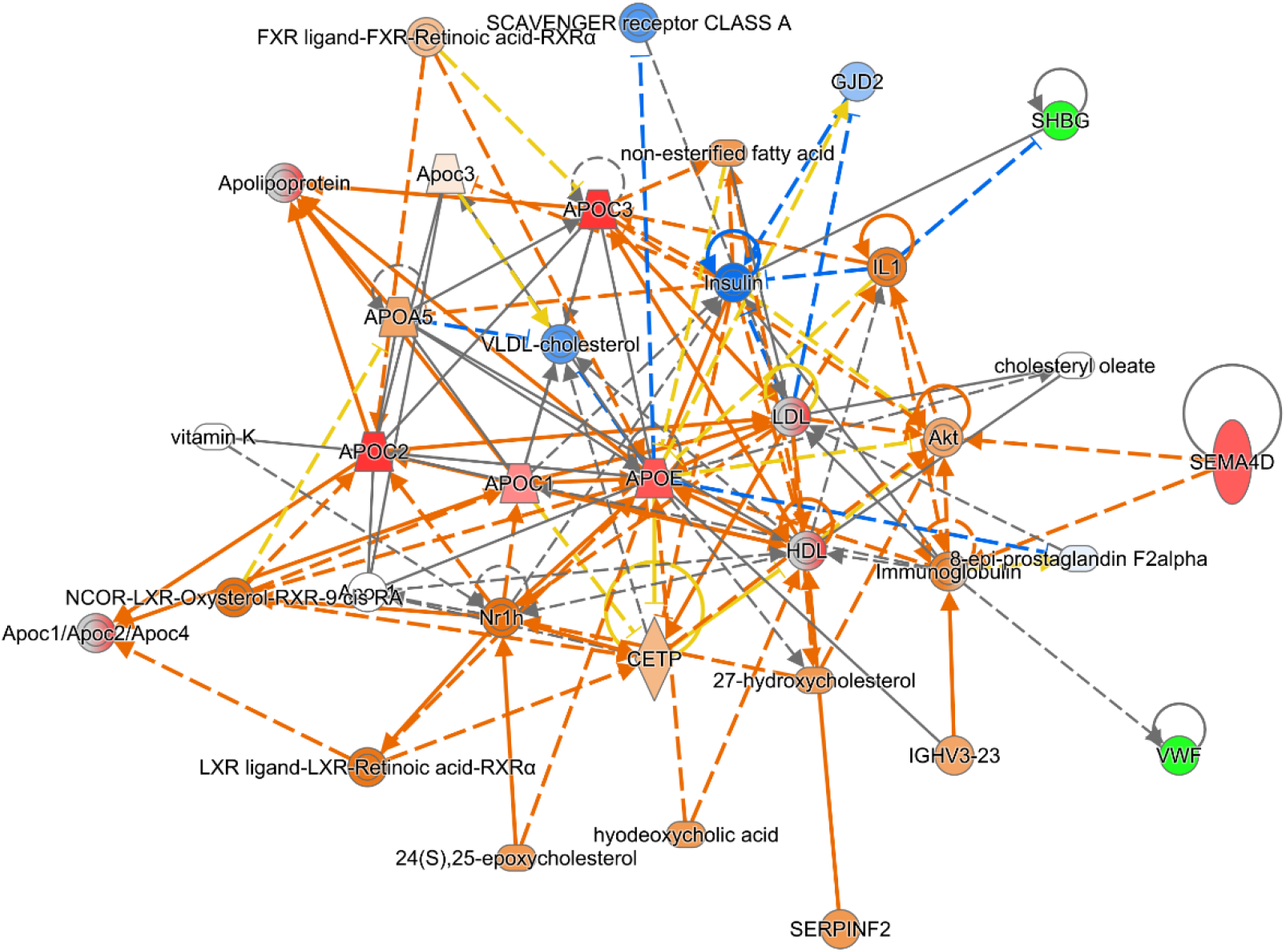
Network enriched using the dysregulated protein from the TG-based MetS patients.

**Figure S3.**
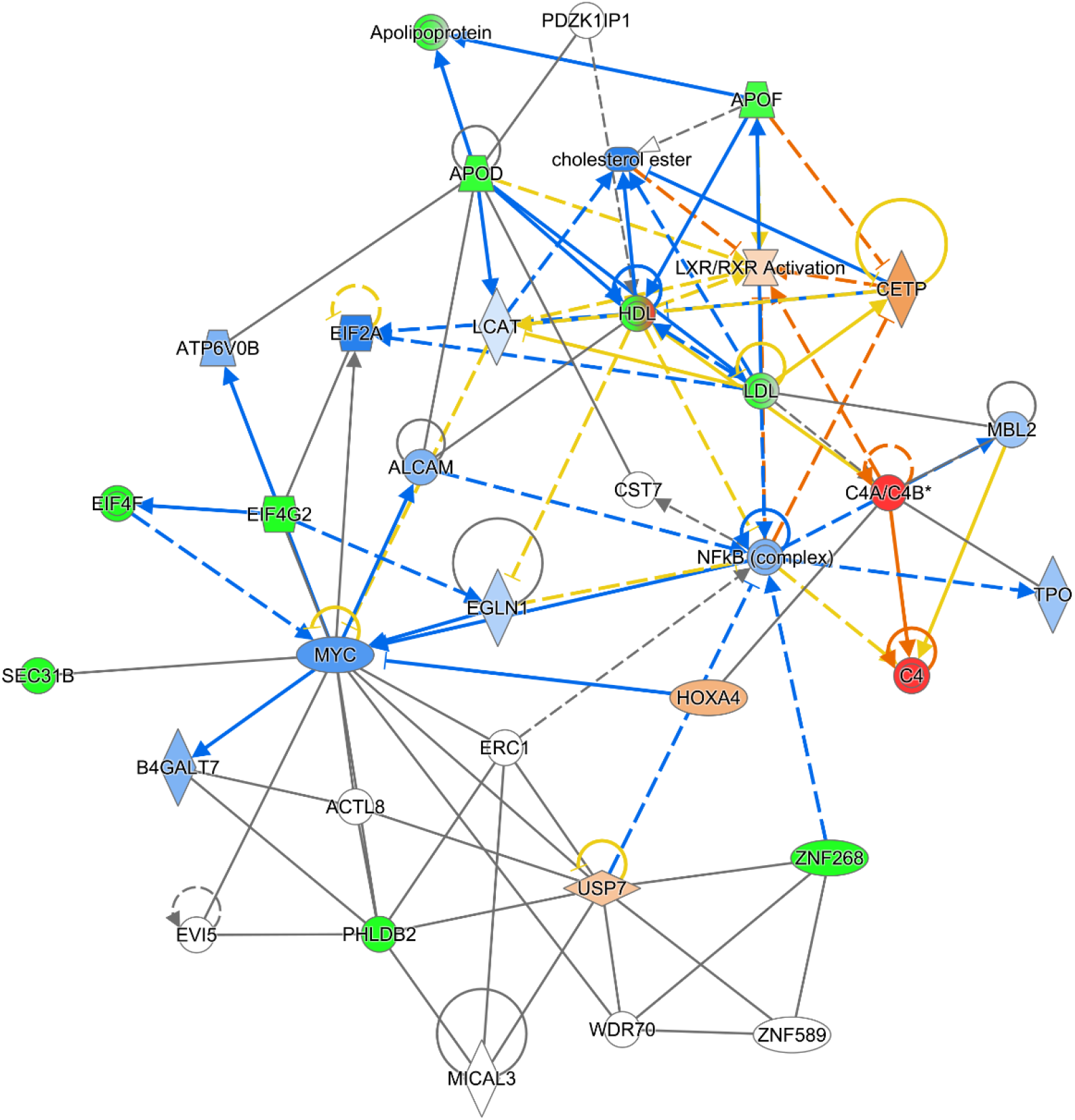
Network enriched using the dysregulated protein from the HDL-based MetS patients.

